# Exploring Intersectionality of Race and Newcomer Status with Material and Social Deprivation in Ontario Census Data: A Comparative Analysis of the ON-MARG Deprivation Index and Machine-Learning Derived Demographic Clusters

**DOI:** 10.1101/2024.10.30.24316442

**Authors:** Renzo Calderon Anyosa, Geoffrey Anderson

## Abstract

**Background:** The Ontario Marginalization Index (ON-MARG) is widely used to assess health inequalities in Ontario by measuring four dimensions of marginalization at the dissemination area (DA) level. However, averaging these dimensions into an overall deprivation score can obscure important information, in particular information on intersectionality of material and social deprivation and race and immigrant status.

**Objective:** To use machine learning algorithms to uncover relationships among the four ON-MARG dimensions across DAs as demographic clusters and to compare the use of these clusters to understand and map marginalization and to describe health inequities.

**Methods:** We applied K-means clustering to 2021 ON-MARG data on the four On-MARG dimensions — Households and Dwellings (HD), Material Resources (MR), Age and Labour Force (AL), and Racialized and Newcomer Populations (RN) across 20,123 DAs. We then compared these clusters to ON-MARG average index scores in terms of mapping marginalization in Toronto and examined how these clusters were associated with inequities in mental health service as compared specific dimensions of the ON-MARG index.

**Results:** We identified four clusters: (1) Advantaged White Canadians, (2) Disadvantaged White Canadians, (3) Advantaged Visible Minorities and Immigrants, and (4) Disadvantaged Visible Minorities and Immigrants. The clustering approach revealed nuanced patterns not captured by the ON-MARG summary scores alone. Disadvantaged White Canadians exhibited the highest outpatient mental health visit rates, particularly among females (250–300 visits per 100,000). Disadvantaged Visible Minorities and Immigrants followed with elevated rates, while both advantaged clusters showed significantly lower utilization. The clusters provided better discrimination of health service disparities than ON-MARG quintiles alone, highlighting that disadvantaged groups, regardless of racial composition, had higher rates of mental health service use.

**Conclusions:** Combining ON-MARG with machine learning clustering offers a more comprehensive understanding of marginalization’s intersectionality, revealing disparities in health service utilization not apparent from the index alone. This approach underscores the need for targeted, intersectional policies to address the specific needs of diverse populations, ultimately contributing to more equitable healthcare interventions in Ontario.

## Introduction

The Ontario Marginalization Index (ON-MARG) is an area-based measure widely used in demographic analysis to understand health inequalities in Ontario (1). Developed using census data, ON-MARG assesses four key dimensions of marginalization: material deprivation, residential instability, dependency, and ethnic concentration (2). It has been instrumental in identifying disparities in health outcomes among different populations within the province, revealing significant differences in rates of overall mortality, mental health issues, and other health-related outcomes across varying levels of deprivation (2–5).

ON-MARG is founded on four statistically derived dimensions, each based on specific census variables:

1. **Households and Dwellings (HD):** Reflecting factors such as living alone or in small family units, residing in rental apartments, and recent residential mobility within the last five years.
2. **Material Resources (MR):** Encompassing indicators like low educational attainment, low-income status, unemployment, single-parent families, reliance on government income support, and poor-quality housing.
3. **Age and Labour Force (AL):** Considering the proportion of younger and older individuals in the neighborhood and the proportion of adults who are unemployed and not seeking employment.
4. **Racialized and Newcomer Populations (RN):** Based on the proportion of people identifying as visible minorities or recent immigrants to Canada within the last five years.

ON-MARG scores are calculated at the dissemination area (DA) level—the smallest census-based geographic unit—each comprising approximately 400 to 700 individuals in stable, cohesive neighborhoods. Higher proportions of the population experiencing specific characteristics (e.g., low income or recent relocation) result in higher deprivation scores. Each dimension provides a numerical value for every DA, facilitating detailed analyses of the relationship between deprivation and health at a granular level.

In practice, ON-MARG can be applied using population-weighted quintiles. DAs are ranked according to their scores on each dimension and divided into quintiles, enabling straightforward comparisons across segments of the population and the creation of maps illustrating the geographic distribution of deprivation. The published ON-MARG includes both dimension scores and quintiles for each DA in Ontario. An overall deprivation quintile for a DA is calculated by averaging the quintiles across all four dimensions.

While ON-MARG has been valuable in highlighting patterns of marginalization, it has limitations, particularly regarding visible minorities and immigrants and the intersection of material and social aspects of deprivation. Developed over a decade ago using data from the 2006 and 2011 censuses, the index’s assumption that a higher proportion of visible minorities and immigrants indicates a specific dimension of marginalization may be less relevant today. Moreover, averaging the quintiles across the four dimensions to derive an overall deprivation score can obscure fundamental differences between neighborhoods. For instance, a neighborhood predominantly composed of visible minorities with median levels of physical and social deprivation would receive the same overall quintile ranking as a predominantly white neighborhood with the highest levels of material and social deprivation. This averaging effect underscores the importance of examining each ON-MARG dimension individually to capture the nuanced realities of marginalized populations.

However, analyzing dimensions separately poses challenges in incorporating the concept of intersectionality. Intersectionality refers to the complex, cumulative ways in which the effects of being a member of a visible minority or a recent immigrant—factors that may result in racism or marginalization—intersect with social and material deprivation (6). In the context of ON-MARG, understanding intersectionality is essential for comprehending how race, ethnicity, and socioeconomic status interact to shape diverse lived experiences and health inequities. Studies have shown that factors such as race, poverty, and lack of social support do not operate in isolation but intersect to create unique disadvantages that significantly impact health outcomes (6–8). An intersectional approach allows for a more nuanced understanding of how marginalization affects different demographic groups.

Studying intersectionality and health inequity involves defining and quantifying the overlaps between visible minority or immigrant status and key dimensions of material and social deprivation within neighborhoods. Identifying distinct groups of neighborhoods that share similar patterns in the distribution of these factors provides a foundation for studying associations between demographic exposures and health outcomes, thereby determining measurable health inequities.

Machine learning offers powerful tools for advancing the study of intersectionality by analyzing complex, multidimensional data. Previous research in the United States has demonstrated that clustering multidimensional demographic features at granular geographic levels, such as block groups and census tracts, can capture socioeconomic gradients in mortality and cancer incidence (9). By leveraging such granular data, machine learning models can uncover nuanced patterns of marginalization, offering a more accurate picture of how different populations experience disparities (10–12). These advances enable researchers to better understand and address the compounded effects of marginalization across intersecting social dimensions.

In this study, we integrate the ON-MARG index with machine learning techniques to analyze demographic data, focusing on how different populations experience and are geographically distributed in terms of marginalization. While ON-MARG provides a useful average index, our aim is to uncover the unique ways in which marginalization manifests within demographic clusters, particularly concerning racialized groups. By employing machine learning, we refine our understanding of how advantaged and disadvantaged populations are distributed across these clusters. To evaluate these differences, we use mental health hospitalizations and outpatient visits as a case study, demonstrating the potential of this approach in assessing healthcare service utilization.

## Methods

### Data Sources

This study utilized the 2021 Ontario Marginalization Index (ON-MARG) data at the Dissemination Area (DA) level, in conjunction with data from the 2021 Canadian Census for the Province of Ontario. ON-MARG measures four key dimensions of marginalization: Households and Dwellings (HD), Material Resources (MR), Age and Labour Force (AL), and Racialized and Newcomer Populations (RN). A detailed list of the indicators for each dimension is provided in the Appendix.

The values for each dimension are represented as factor scores, which are derived from principal component factor analysis. These scores are on an interval scale, standardized with a mean of zero and a standard deviation of one. Lower factor scores indicate areas with the least marginalization, while higher scores correspond to areas with the most marginalization. Additionally, the data is presented in quintiles, created by ranking the factor scores and dividing the geographic units into five groups, each representing 20% of the population. These quintiles range from 1 (least marginalized) to 5 (most marginalized) (1).

In addition to the factor scores and quintiles, we also created deciles based on the factor scores. These deciles were generated using the same methodology as the quintiles but divide the geographic units into groups representing 10% of the population each. This approach was implemented to capture more granular differences among groups and to provide a larger range of values for use in the clustering algorithm.

Census variables corresponding to each ON-MARG dimension were also identified to replicate the ON-MARG indicators to assess the distribution of each indicator beyond the index itself (Appendix 1).

Healthcare data was obtained from the Ontario Health Data Platform (OHDP-Q), a comprehensive resource that integrates various health databases from Ontario. For this study, we utilized data from 2021 to 2023, specifically focusing on hospitalization rates from the Discharge Abstract Database (DAD) and outpatient visits from the National Ambulatory Care Reporting System (NACRS). We selected ICD 10 codes related to mental health cases based on the indicator creation guidelines from the Mental Health and Addictions System Performance in Ontario. Each hospitalization and visit were linked to the corresponding Dissemination Area using the postal code, allowing for a precise geographical analysis of healthcare utilization across different populations.

### Machine learning clustering analysis

To complement the traditional ON-MARG analysis, machine learning techniques for clustering analysis were used. The primary goal was to group Dissemination Areas (DAs) into distinct clusters based on their marginalization profiles across the four ON-MARG dimensions. K-Means clustering was selected for this purpose, with 300 random starts to ensure robustness and avoid local minima. This method allowed us to identify the model that best minimized within-cluster distances while maximizing the distances between clusters.

The deciles of each of the four ON-MARG dimensions were used as the primary input, providing a standardized representation of the marginalization index that accounts for the population distribution within each DA. This approach ensured that the clustering was reflective of the underlying population dynamics across all dimensions. Additionally, as a sensitivity analysis, the raw index values were also used as input to the K-Means algorithm to further evaluate the stability and robustness of the clustering outcomes.

The optimal number of clusters was determined using the elbow method, which identifies the point at which adding additional clusters no longer significantly reduces the within-cluster sum of squares. This method allowed us to balance the complexity of the model with the interpretability of the clusters.

### Statistical Analysis

We compared the composition of each cluster, with a particular emphasis on the mean proportions of each ON-MARG indicator within the clusters. The analysis aimed to uncover patterns of marginalization by evaluating the mean ON-MARG scores across clusters.

Given that the majority of ON-MARG indicators are derived from population or dwelling proportions, the analysis calculated the mean and 95% confidence intervals (CI) for the proportion of each indicator across all DAs within each cluster. This provided measure of central tendency and variability within each cluster.

To assess differences between clusters, we calculated the mean pairwise differences and their corresponding 95% confidence intervals (CIs) for each ON-MARG indicator. This analysis helped us identify statistically significant differences between clusters, highlighting where marginalization profiles diverge.

We also examined the distribution of the recommended ON-MARG summary score to see how it varies across clusters. For evaluating healthcare service use, we calculated monthly hospitalization and outpatient visit rates per 100,000 people for each cluster, stratified by sex. Additionally, we computed overall rate ratios adjusted for calendar month to assess differences between clusters. All statistical analyses were performed using R version 4.3.

## Results

### Cluster Analysis

The clustering analysis identified four distinct profiles of Dissemination Areas (DAs) based on their marginalization characteristics (Elbow method shown in Appendix Figure 1). The mean decile scores for each ON-MARG dimension across these clusters are summarized in Figure 1. Each cluster is described below, accompanied by a label that encapsulates the general characteristics of its population. These labels serve as summaries and may not precisely represent every individual within the DAs.

**Figure 1:**
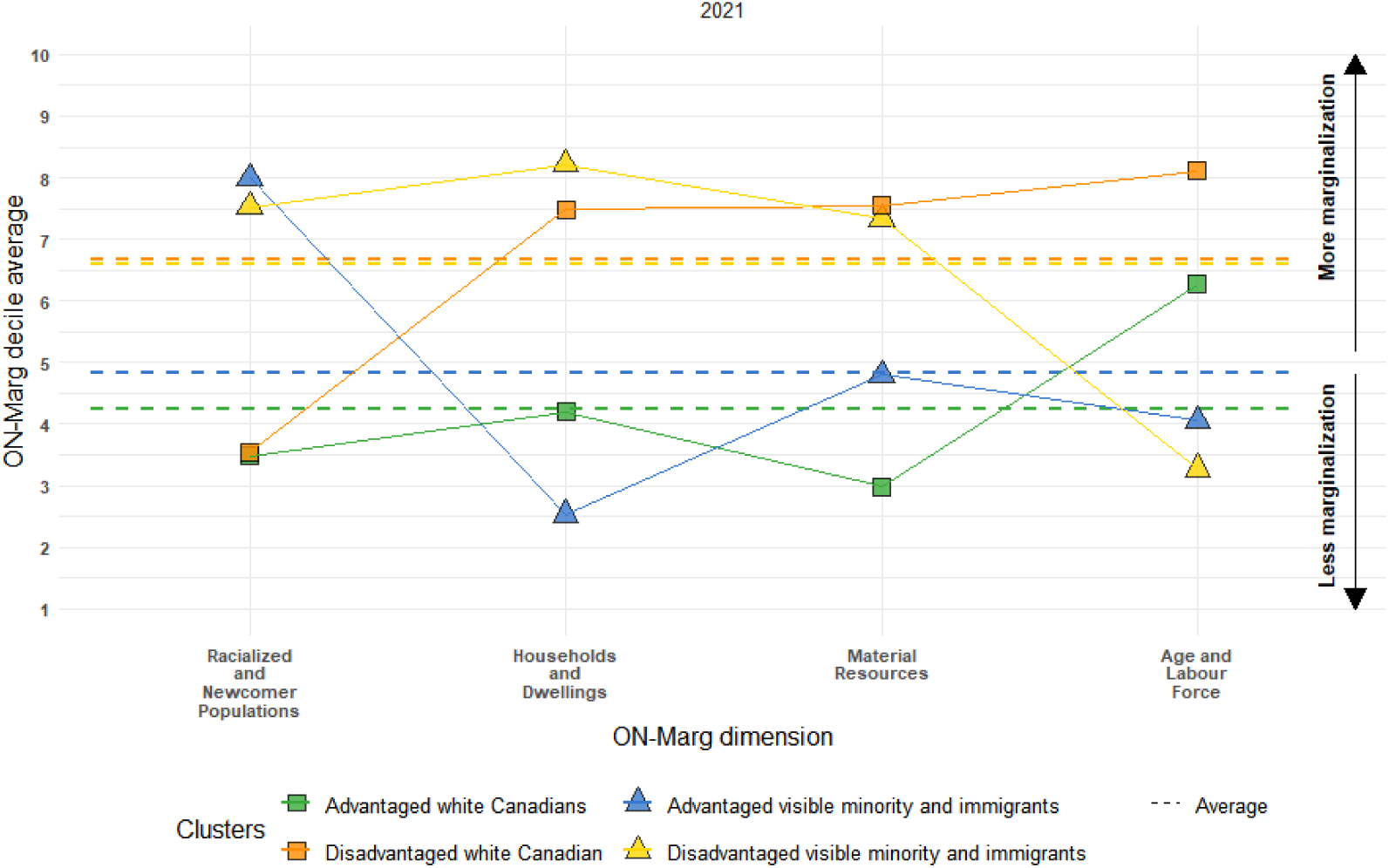
Average ON-MARG Deciles Across Clusters for 2021 by ON-MARG Dimension.

**Figure 2.**
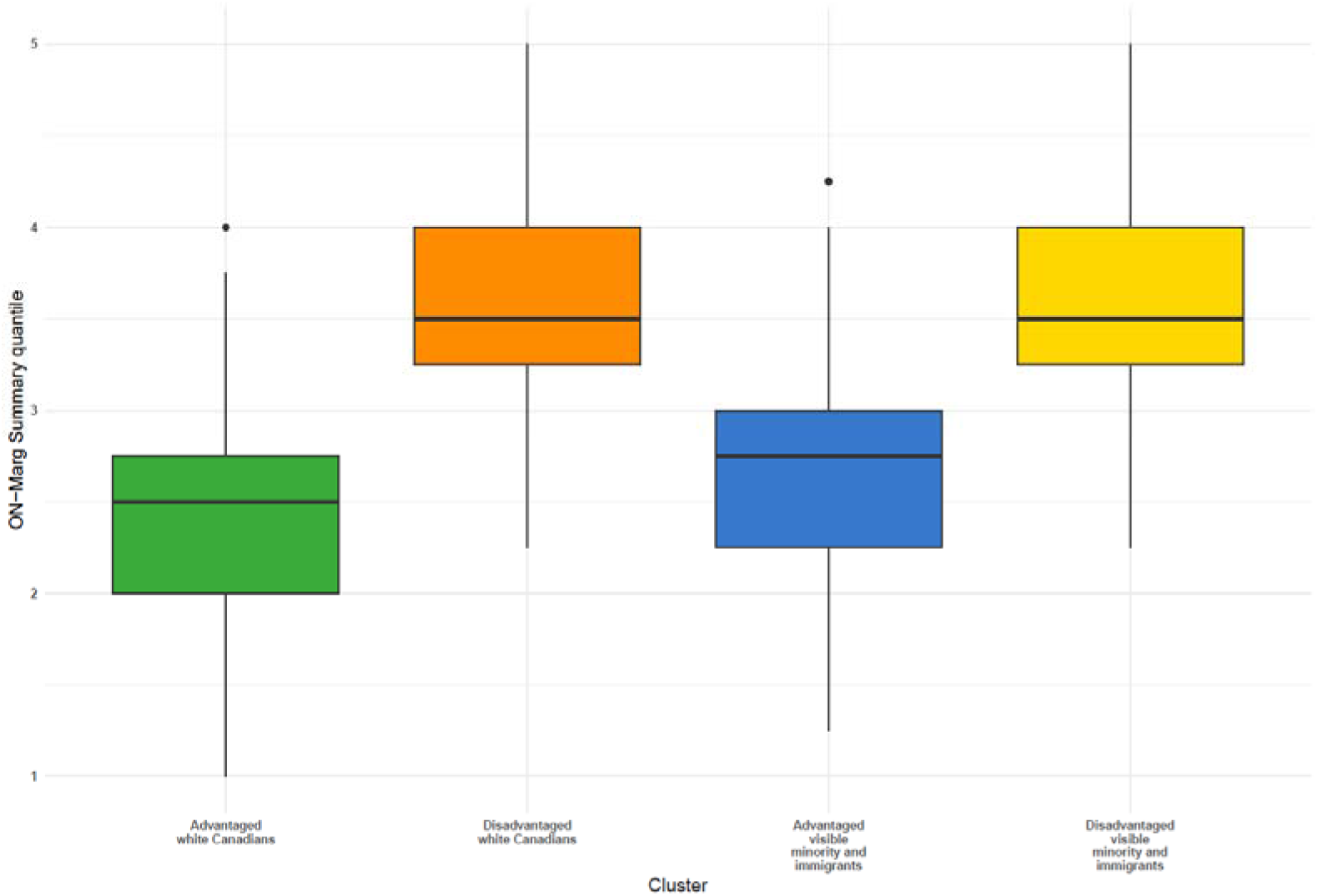
Summary Score for the ON-MARG dimensions by Clusters.

#### Cluster 1: Advantaged White Canadians

This cluster comprises DAs with low proportions of visible minorities and immigrants (average 3rd decile in the RN dimension), indicating a higher presence of white Canadians. Residents here are relatively advantaged, scoring below the mean in the Households and Dwellings (HD) dimension (average 4th decile) and the Material Resources (MR) dimension (average 3rd decile). They score slightly above average in the Age and Labour Force (AL) dimension (average 6th decile).

#### Cluster 2: Disadvantaged White Canadians

Similar to Cluster 1 in demographic composition (average 3rd decile in the RN dimension), this cluster includes DAs predominantly inhabited by white Canadians. However, these areas are relatively deprived, with high scores in the HD (average 7th decile) and MR (average 7th decile) dimensions, and a very high score in the AL dimension (average 8th decile).

#### Cluster 3: Advantaged Visible Minorities and Immigrants

This cluster consists of DAs with high proportions of racialized and newcomer populations (average 7th decile in the RN dimension). Residents are relatively advantaged, exhibiting low levels of deprivation in the HD (average 2nd decile), MR (average 4th decile), and AL (average 3rd decile) dimensions.

#### Cluster 4: Disadvantaged Visible Minorities and Immigrants

Also characterized by high proportions of racialized and newcomer populations (average 7th decile in the RN dimension), this cluster differs from Cluster 3 by displaying high levels of deprivation in the HD (average 8th decile) and MR (average 7th decile) dimensions, while maintaining similar levels in the AL dimension (average 3rd decile).

These clusters highlight the importance of intersectionality in understanding Ontario’s demographics. Neighborhoods predominantly inhabited by racialized individuals or newcomers, as well as those predominantly inhabited by white Canadians, can each be subdivided into relatively advantaged and disadvantaged groups based on other dimensions of marginalization. This nuanced perspective differs from that provided by the overall ON-MARG deprivation score, which averages across all dimensions. For instance, as shown in Table 1, Clusters 2 and 4 differ significantly in race and immigrant status but have similar overall ON-MARG quintile scores; the same pattern is observed for Clusters 1 and 3.

**Table 1.** Decile average distribution across clusters and ON-MARG dimensions.

|  | Cluster 1:<br>Advantaged<br>white<br>Canadians | Cluster 2:<br>Disadvantaged<br>white<br>Canadian | Cluster 3:<br>Advantaged<br>visible minority<br>and immigrants | Cluster 4:<br>Disadvantaged<br>visible minority and<br>immigrants |
| --- | --- | --- | --- | --- |
| Households and Dwellings | 4.2 | 7.5 | 2.5 | 8.2 |
| Material Resources | 3.0 | 7.6 | 4.8 | 7.3 |
| Age and Labour Force | 6.3 | 8.1 | 4.1 | 3.3 |
| Racialized and Newcomer Populations | 3.5 | 3.5 | 8.0 | 7.5 |
| **Overall decile average** | **4.2** | **6.7** | **4.8** | **6.5** |
| **Overall quintile average** | **2.4** | **3.6** | **2.7** | **3.5** |

The clustering approach allows for the assignment of every Ontario DA to a specific cluster. As detailed in Table 2, the four clusters collectively encompass the entire Ontario population, each accounting for approximately one-quarter of the total population.

**Table 2.** Distribution of Mean Population per Dissemination Area (DA), Total Population, and Total DAs by Cluster for.

| Cluster | Mean population per DA | Total population | Total DAs |
| --- | --- | --- | --- |
| 1. Advantaged white Canadians | 584.5 | 3452180 (24%) | 5906 (29%) |
| 2. Disadvantaged white Canadian | 584.1 | 2813166 (20%) | 4816 (24%) |
| 3. Advantaged visible minority and immigrants | 864.8 | 4171955 (29%) | 4824 (24%) |
| 4. Disadvantaged visible minority and immigrants | 814.5 | 3727925 (26%) | 4577 (23%) |
| Total | 712.0 | 14165226 | 20123 |

To compare the clusters with the ON-MARG summary scores, we categorized the summary scores into four groups: less than 2, between 2 and 3, between 3 and 4, and 4 or higher. An examination of the distribution of clusters within these summary score groups revealed notable trends (Figure 3). In the low marginalization group (summary score < 2), 67% of DAs belonged to the Advantaged White Canadians cluster, while 33% were part of the Advantaged Visible Minorities and Immigrants cluster. In the group with summary scores between 2 and 3, the distribution was 54% Advantaged White Canadians, 34% Advantaged Visible Minorities and Immigrants, 5% Disadvantaged White Canadians, and 7% Disadvantaged Visible Minorities and Immigrants.

**Figure 3.**
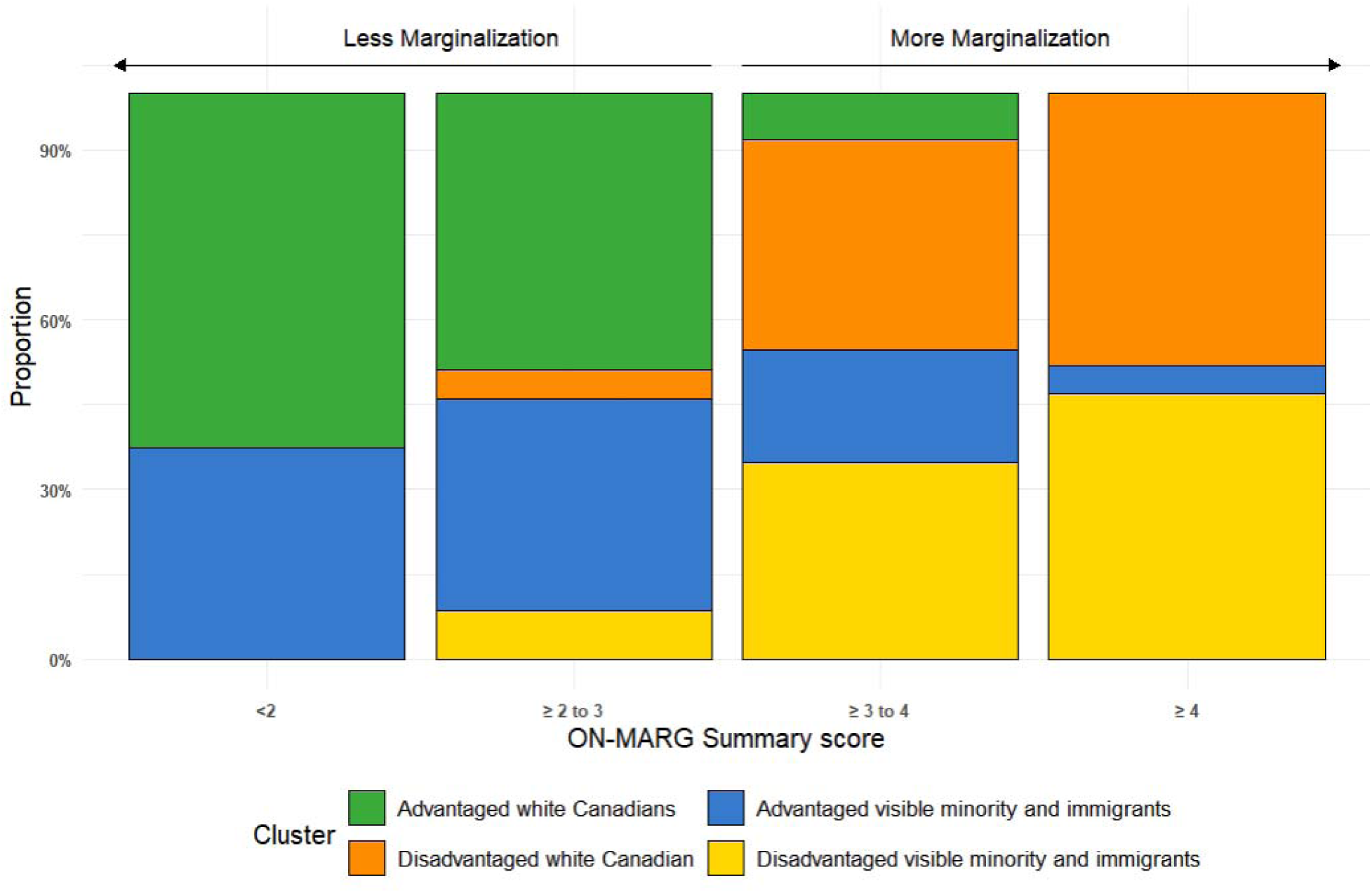
Cluster Distribution Across Different Levels of ON-MARG Summary Scores.

As marginalization increased (summary scores between 3 and 4), the distribution shifted: 37% Disadvantaged White Canadians, 33% Disadvantaged Visible Minorities and Immigrants, 10% Advantaged White Canadians, and 20% Advantaged Visible Minorities and Immigrants. In the highest marginalization group (summary score ≥ 4), 49% were Disadvantaged White Canadians, 46% were Disadvantaged Visible Minorities and Immigrants, and only 5% were Advantaged Visible Minorities and Immigrants.

Spatial analysis of the Toronto Census Metropolitan Area provided a visual comparison of the geographic distribution of clusters and ON-MARG summary score categories (Figure 4). The cluster map revealed distinct geographic patterns, with certain clusters more prevalent in specific areas. For instance, Advantaged White Canadians dominated central and some western regions, while Disadvantaged Visible Minorities and Immigrants were concentrated in the northwest and northeast.

**Figure 4.**
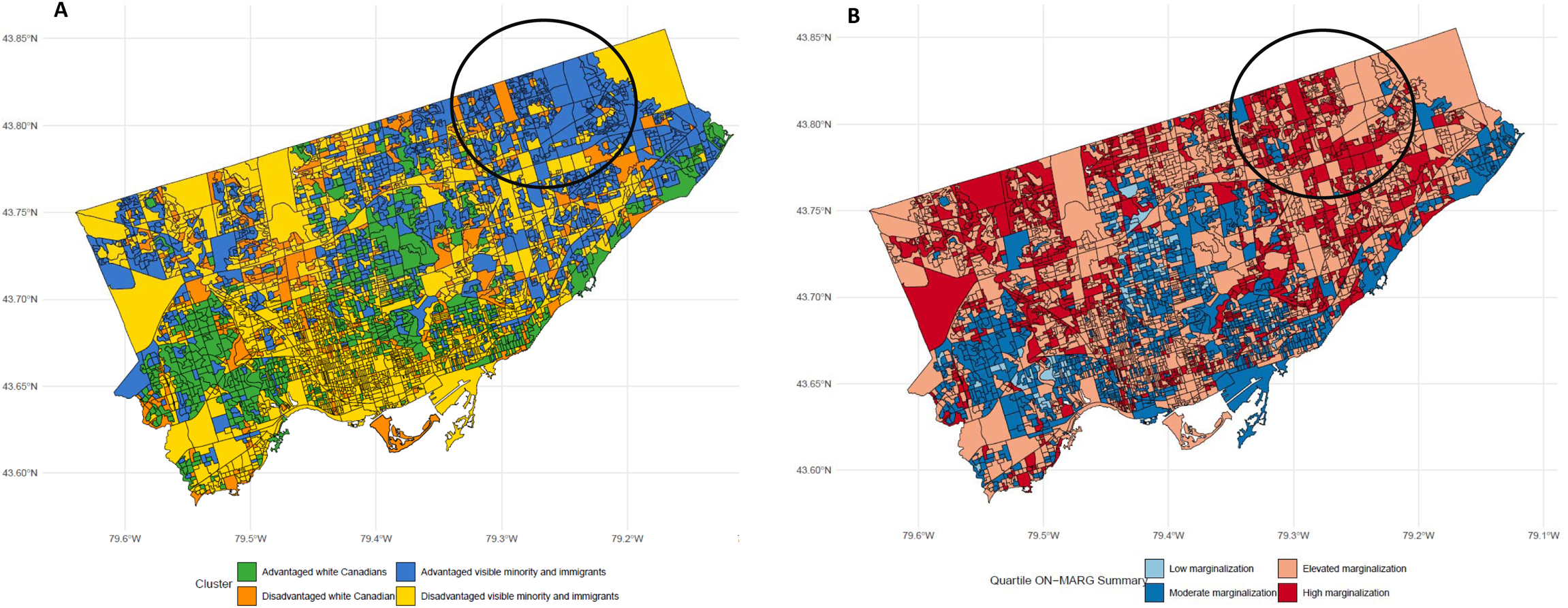
Comparison of ON-Marg summary score and clusters geographical distribution.

The ON-MARG summary score map highlighted varying degrees of marginalization across the region but did not fully capture nuances within the Advantaged Visible Minorities and Immigrants group. For example, the northeastern area, primarily composed of Advantaged Visible Minorities and Immigrants, was categorized as having elevated or high marginalization according to the summary score. This spatial analysis underscores the complex intersection of race, socioeconomic status, and geographic location, offering deeper insights into structural inequities.

### Healthcare Utilization Analysis

To assess how the clusters and the ON-MARG overall score describe and explain health inequities, we analyzed outpatient physician services for non-psychotic mental health diagnoses at the DA level from January 2021 through January 2023.

### Outpatient Visit Rates by Cluster

Outpatients visit rates for mental health conditions varied significantly among the clusters (Figure 5). The Disadvantaged White Canadians cluster exhibited the highest rates, particularly among females, ranging from 250 to 300 visits per 100,000 population. Males in this cluster also showed elevated rates, generally between 200 and 250 visits per 100,000. The Disadvantaged Visible Minorities and Immigrants cluster followed, with females registering 150 to 200 visits per 100,000 and males ranging from 120 to 170 visits per 100,000.

**Figure 5.**
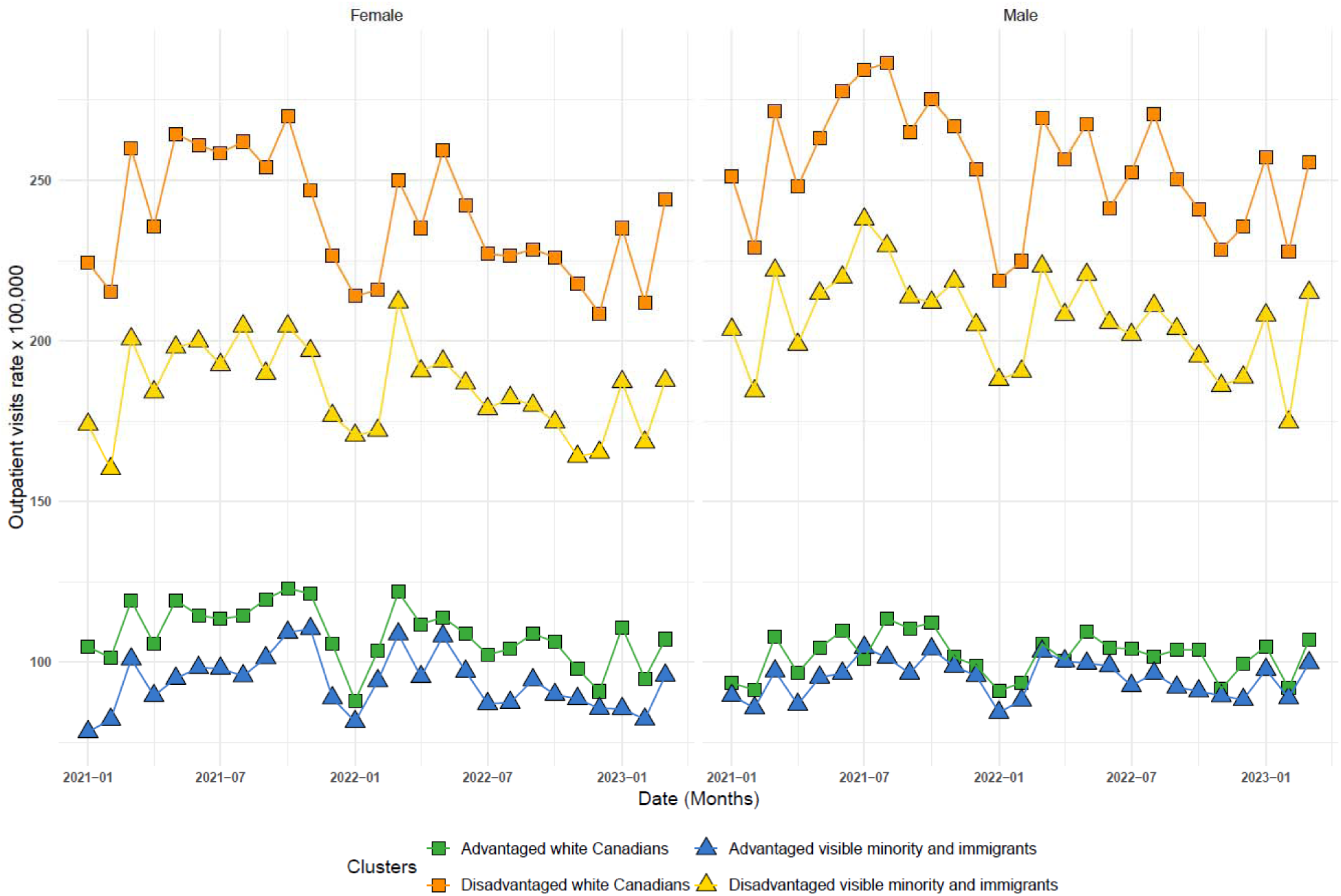
Outpatient visits Rates by Gender and Cluster Over Time (2021-2023)

In contrast, the Advantaged clusters displayed much lower rates. Among males, the lowest rates were observed in the Advantaged White Canadians cluster (approximately 90 visits per 100,000) and the Advantaged Visible Minorities and Immigrants cluster (approximately 70 visits per 100,000). These patterns highlight clear disparities in outpatient visit rates across different demographic clusters.

Rate ratio analysis further emphasized these disparities (Figure 6). Among females, the highest rate ratio was observed when comparing the Disadvantaged White Canadians cluster to the Advantaged Visible Minorities and Immigrants cluster (rate ratio 2.5, 95% CI: 2.4–2.6), mirroring patterns seen in hospitalization data. Although rate ratios for males were slightly higher than for females, significant disparities persisted, particularly when contrasting disadvantaged groups with advantaged ones, especially concerning the visible minority component. These findings underscore ongoing inequalities in healthcare access and outcomes across different demographic groups.

**Figure 6.**
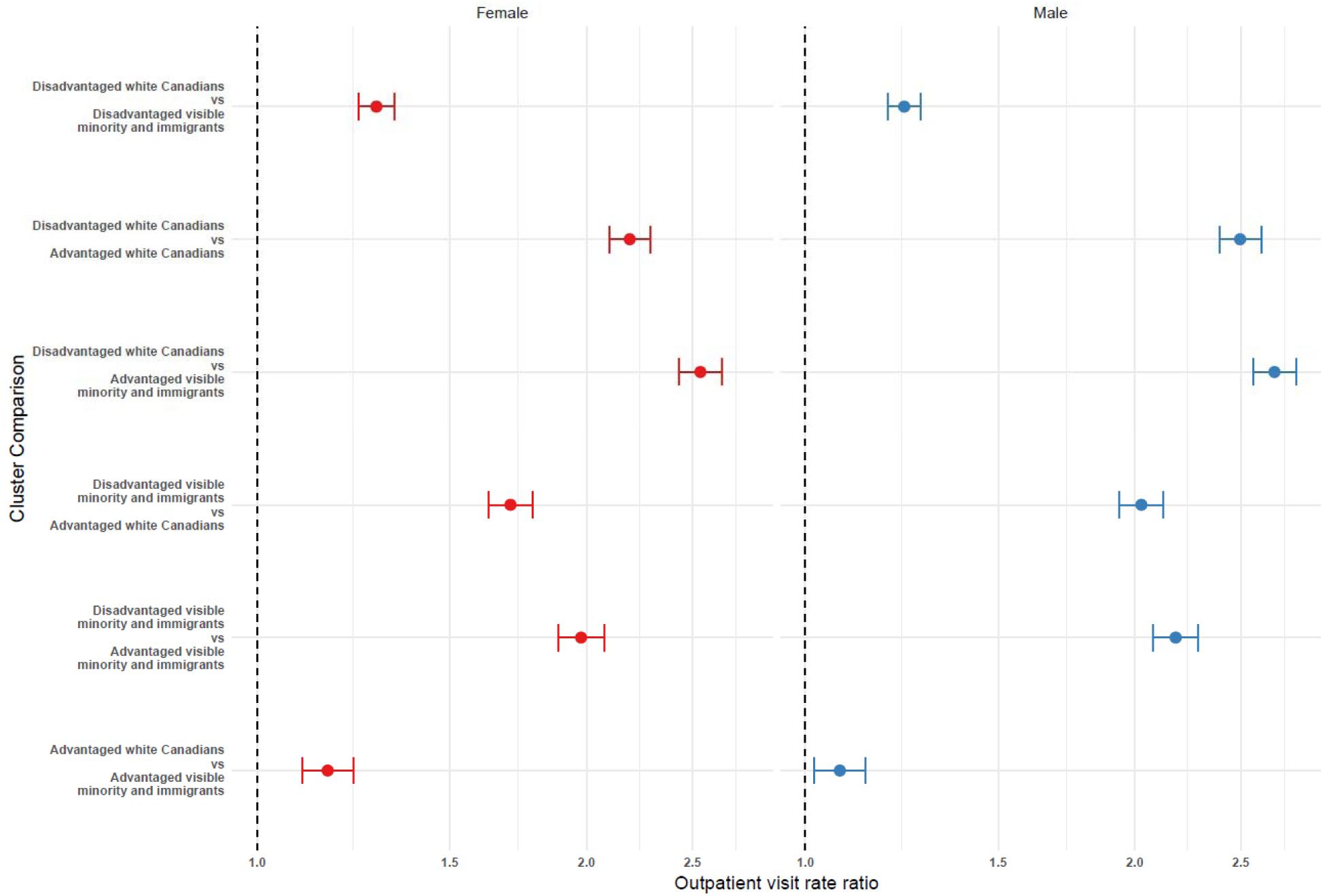
Comparison of Outpatient visits Rate Ratios by Gender and Cluster (2021-2023)

### Outpatient Visit Rates by Material Deprivation Quintile

Analyzing outpatient visit rates by the Material Resources (MR) deprivation quintiles revealed that the highest rates were among individuals in the 5th quintile (most deprived) (Figure 7). Females in this quintile consistently exceeded 250 visits per 100,000, peaking above 300 in some months. Males exhibited similarly elevated rates, though slightly lower than females, ranging between 200 and 250 visits per 100,000. In contrast, both genders in the 1st quintile (least deprived) consistently showed the lowest outpatient visit rates, generally around 100 visits per 100,000. The 4th quintile also demonstrated high rates, particularly among females, while the 2nd and 3rd quintiles maintained intermediate levels of outpatient visits.

**Figure 7.**
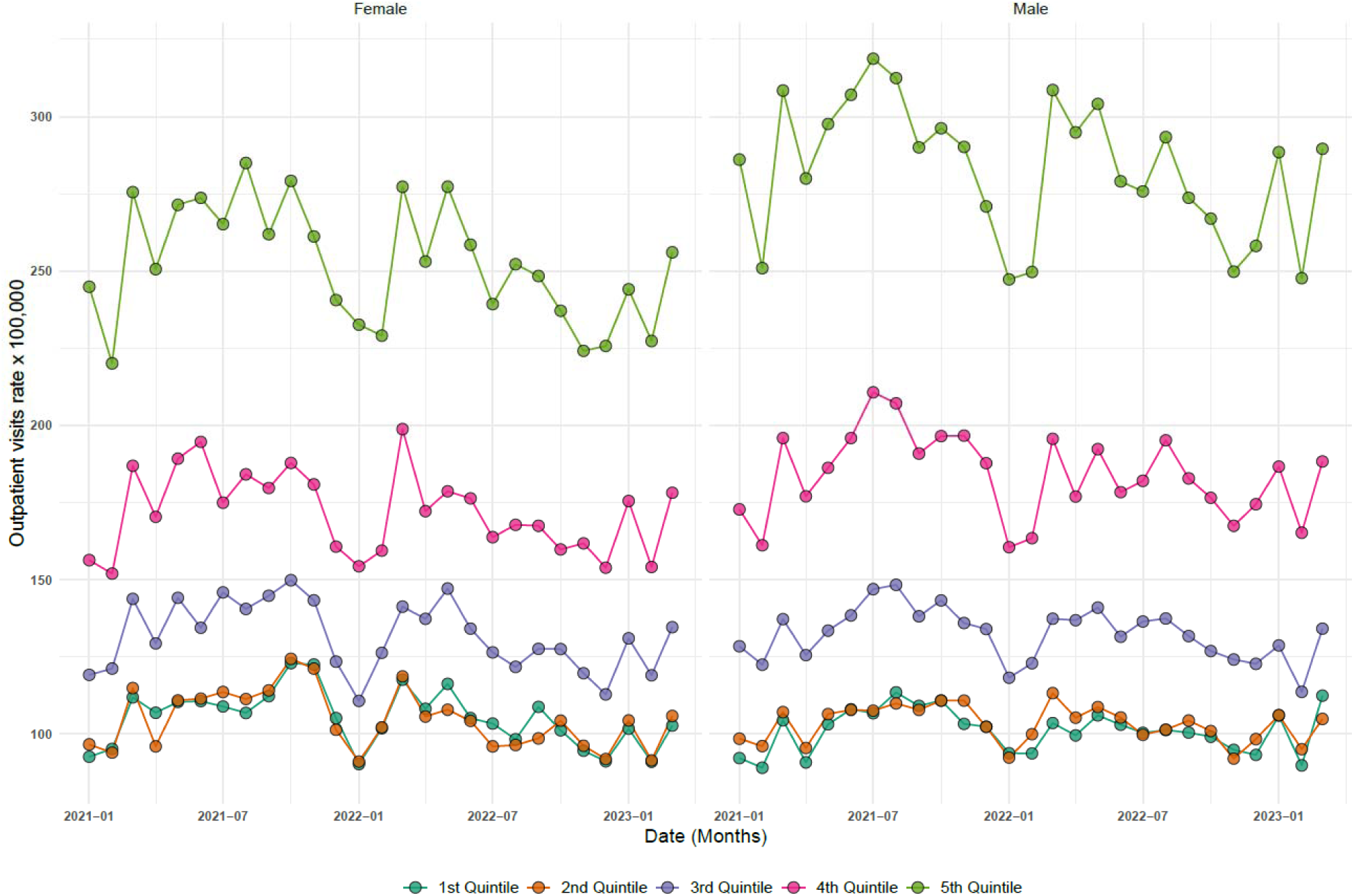
Outpatient visits Rates by Gender and Material Deprivation Quantile Over Time (2021-2023)

### Outpatient Visit Rates by Racialized and Newcomer Population Quintile

When examining outpatient visit rates by the Racialized and Newcomer Populations (RN) deprivation quintiles, both females and males in the 1st quintile (areas with lower proportions of racialized individuals and newcomers) consistently showed the highest rates, ranging from 180 to 210 visits per 100,000 (Figure 8). This indicates greater healthcare utilization among these populations. In contrast, the 5th quintile (areas with higher proportions of racialized individuals and newcomers) exhibited significantly lower rates, generally around 120 visits per 100,000. The intermediate quintiles (2nd, 3rd, and 4th) displayed fluctuating trends, with visit rates between 150 and 180 per 100,000.

**Figure 8.**
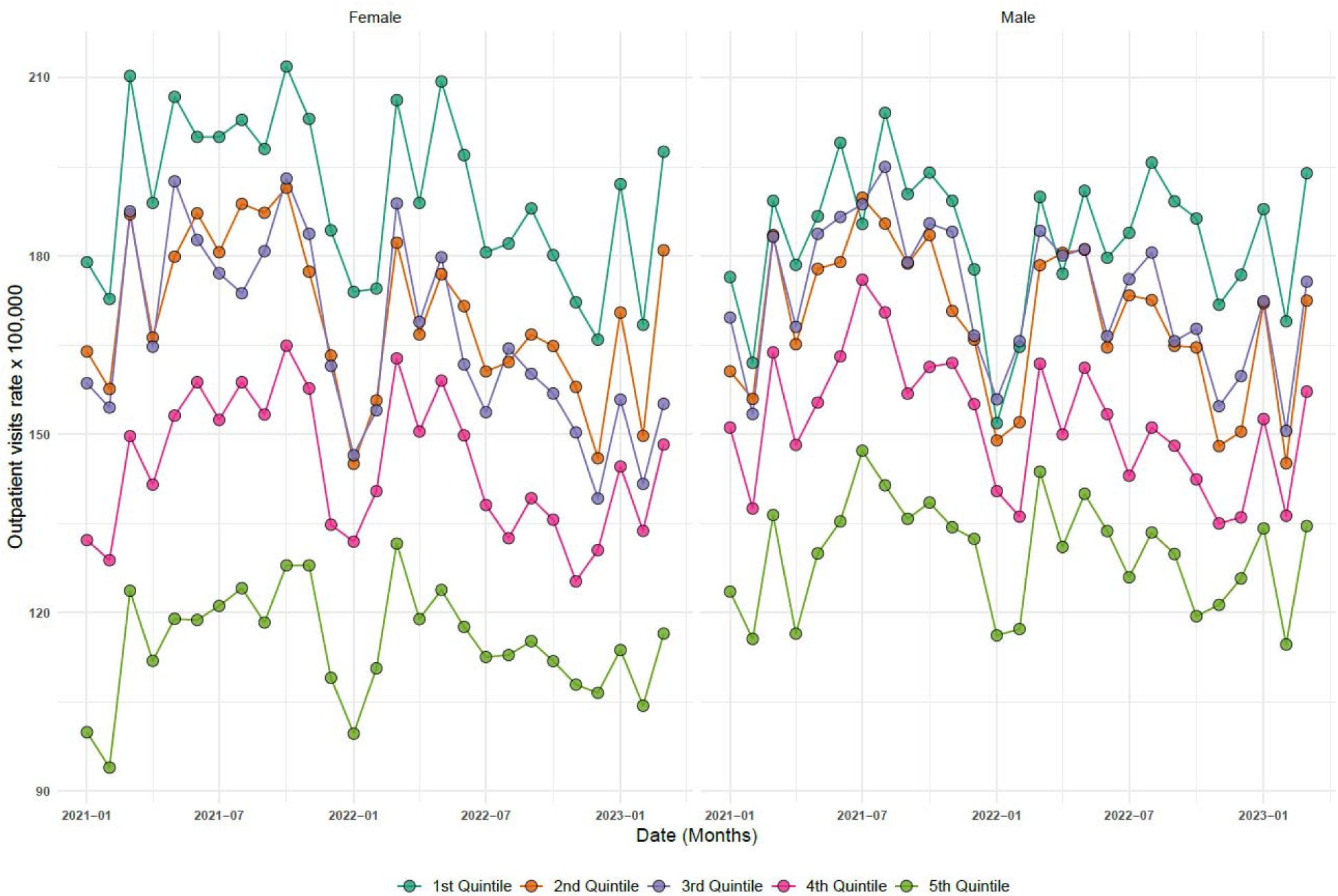
Outpatient visits Rates by Gender and Newcomer Population Quantile Over Time (2021-2023)

By relying solely on the marginalization index quantiles, important nuances were overlooked. The index quantiles did not differentiate between areas with similar deprivation scores but different demographic compositions, nor did they reveal the compounded effects of intersecting marginalization dimensions on healthcare utilization. In contrast, the clustering approach provided a more comprehensive understanding by integrating multiple dimensions of marginalization, leading to more precise identification of populations at risk and the disparities in healthcare service use.

## Discussion

This study provides a novel exploration of the intersectionality present within demographic clusters in Ontario, specifically examining the diverse experiences of marginalization among White Canadians and visible minority and immigrant groups. By integrating the Ontario Marginalization Index (ON-MARG) with machine learning clustering techniques, we have been able to unpack part of the complex interplay between different dimensions of marginalization, offering deeper insights into how these factors interact and manifest across different populations.

Our approach offers a fresh perspective on summarizing ON-MARG by emphasizing the critical role of intersectionality in understanding marginalization. While traditional ON-MARG analysis is valuable, it often obscures key details by averaging out significant features, particularly in how various dimensions of marginalization intersect. For instance, our clustering analysis highlighted that visible minority and immigrant groups face a distinct form of marginalization that cannot be fully understood by examining any single ON-MARG dimension. Relying solely on averages risks overlooking the diverse marginalization profiles that exist within a single summary measure. This underscores the importance of intersectional approaches that recognize and account for the combined effects of these dimensions, rather than treating them in isolation.

The identification of four distinct clusters—Advantaged White Canadians, Disadvantaged White Canadians, Advantaged Visible Minorities and Immigrants, and Disadvantaged Visible Minorities and Immigrants—underscores the varied ways in which marginalization manifests across Ontario. The marked contrasts between the “advantaged” and “disadvantaged” clusters within both White Canadian and visible minority/immigrant groups reveal that socio-economic factors such as material resources and labor force participation are critical determinants of these disparities.

The Disadvantaged Visible Minorities and Immigrants cluster, in particular, not only endures significant racialized marginalization but also grapples with severe household, dwelling, and material deprivation compared to its disadvantaged white Canadian counterparts. While both disadvantaged groups face challenges within their respective racialized categories, the extent of marginalization is more pronounced among visible minorities and immigrants. This heightened marginalization aligns with existing research, which indicates that visible minorities and recent immigrants often reside in more economically deprived urban areas, further exacerbating their material deprivation and restricting their access to well-paying jobs (13).

The residential segregation prevalent in Canada’s largest cities intensifies these disparities, as visible minorities are more likely to be concentrated in neighborhoods with lower socioeconomic status, leading to an increased risk of poverty and social isolation (14). Moreover, systemic discrimination in housing and employment markets compounds these challenges, resulting in overcrowded living conditions and limited economic mobility for visible minority groups (15). The intersection of racialized identity with economic disadvantage not only perpetuates material deprivation but also has significant implications for health outcomes. Visible minorities in deprived neighborhoods are more likely to experience poorer health, driven by the chronic stress and social isolation associated with their marginalized status (16).

The pronounced gaps in homeownership and the prevalence of apartment living between the disadvantaged visible minority and immigrant group and their white counterparts highlight how housing inequities persist as a significant barrier to economic stability and social mobility. Moreover, the higher proportion of recent immigrants and visible minorities in these disadvantaged clusters further complicates their economic integration, as these populations face additional challenges related to language barriers, cultural adjustment, and discrimination in the labor market. Language proficiency, in particular, remains a formidable obstacle for recent immigrants, impeding their social integration and professional advancement. The challenge is exacerbated by accent discrimination, which not only limits employment opportunities but also reinforces social exclusion within the workplace(17). This exclusion extends beyond economic outcomes, significantly affecting the mental and physical well-being of immigrants, especially visible minorities who experience daily microaggressions and systemic barriers. The cumulative stress of these experiences contributes to poorer health outcomes, further marginalizing these groups and perpetuating the cycle of disadvantage (18).

When evaluating healthcare outcomes, the clusters revealed distinct profiles that show the diverse challenges faced by each group. Our analysis showed that the White Canadian group exhibited higher levels of hospitalization and outpatient visits. This pattern could be influenced by several factors, including the “healthy immigrant effect” (HIE), where recent immigrants generally arrive in better health. Studies in Canada have demonstrated that this effect can mask underlying health issues, particularly in mental health, where racialized immigrants are more likely to have undiagnosed or unrecognized conditions like late-life depression (19,20). This could partly explain why visible minority populations, despite facing significant barriers to healthcare access, might report lower hospitalization rates; their health issues may remain undetected or untreated.

Moreover, studies have suggested that the HIE diminishes over time, especially in mental health, which might contribute to the observed differences in healthcare utilization between White Canadians and long-term immigrants (21). Moreover, other studies have support this by showing that both White and visible minority immigrants who have been in Canada for 10 to 20 years exhibit worse mental health outcomes compared to more recent immigrants (22). This could indicate a gradual erosion of the HIE, leading to increased healthcare needs over time.

By highlighting these disparities, the clustering approach provides a clearer understanding of the unique healthcare challenges within each group. It underscores the need for targeted interventions that not only address immediate healthcare access but also consider the long-term health trajectories of immigrant populations, particularly those within visible minority groups, as they navigate the healthcare system in Canada.

The identification of these clusters is crucial for advancing research and informing policy because it moves beyond the limitations of traditional regression adjustments including summary measures that often reduce complex social realities to mere numbers. By breaking down the Ontario Marginalization Index into distinct demographic clusters, this analysis allows researchers to delve deeper into the lived experiences of different communities, recognizing that marginalization is not a uniform experience but varies significantly across populations.

For instance, the Disadvantaged Visible Minorities and Immigrants cluster not only faces racialized marginalization, but also distinct challenges related to housing, employment, and health that are not as prevalent in the Disadvantaged White Canadians cluster. Understanding these nuances enables a more targeted approach to interventions, whether in public health, social services, or housing policy. For example, tailored health programs that address the specific stressors and health disparities faced by visible minorities in deprived neighborhoods can be developed, rather than relying on a one-size-fits-all approach.

Moreover, this clustering approach can help identify communities that are particularly vulnerable to intersecting forms of disadvantage, allowing for more effective allocation of resources and support. For example, in urban planning, recognizing the concentration of marginalized groups in specific areas can lead to more inclusive development strategies that address both economic and social inequities. This shift from a broad, generalized understanding of marginalization to a more detailed, community-focused perspective can lead to more equitable outcomes and a better understanding of the unique challenges faced by different demographic groups.

Despite the strengths of this study, there are limitations that should be acknowledged. The reliance on ON-MARG and census data, while comprehensive, may not capture all relevant dimensions of marginalization, such as those related to health or social capital. Additionally, the clustering approach, while robust, is still subject to the inherent biases and assumptions of the algorithms used. Future research could explore the integration of other data sources, such as health records or social surveys, to provide an even richer understanding of marginalization.

Moreover, longitudinal studies that track changes in these clusters over time would be valuable in assessing the long-term impacts of marginalization and the effectiveness of policy interventions. Understanding how these clusters evolve could offer insights into the dynamics of socio-economic mobility and the persistence of inequalities.

## Conclusion

This study highlights the utility of combining traditional indices like ON-MARG with advanced machine learning techniques to explore the intersectionality of marginalization. By identifying distinct clusters of White Canadians and visible minority/immigrant groups, we have provided a clearer picture of how different forms of marginalization intersect and manifest across Ontario. These insights highlight the need for targeted, intersectional policy approaches that address the specific needs of diverse populations, ultimately contributing to more equitable and effective interventions.

## Data Availability

CENSUS data and ON-Marg data are publicly available, health care data is available trough the Ontario Health Data Platform.

## Appendix

**Appendix Figure 1.**
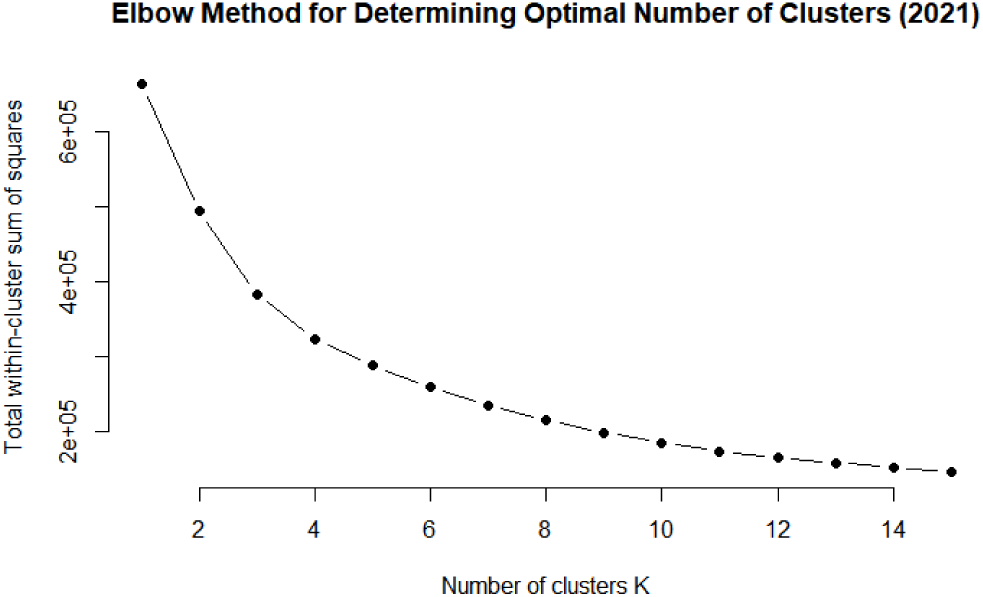
Elbow method for determining the optimal number of clusters.

Appendix List 1. ON-MARG dimensions and indicators from the Census

Households and Dwellings Indicators

1. Proportion of the population living alone
2. Proportion of the population who are not in the youth age group (ages 5-15)
3. Average number of people per dwelling
4. Proportion of dwellings that are apartment buildings
5. Proportion of the population who are single, divorced, or widowed
6. Proportion of dwellings that are rented (not owned)
7. Proportion of the population who moved within the past five years

Material Resources Indicators

1. Proportion of the population aged 25 to 64 without a high school diploma
2. Proportion of families that are single-parent households
3. Proportion of total income coming from government transfers
4. Proportion of the population aged 15 and older who are unemployed
5. Proportion of the population living below the low-income threshold
6. Proportion of households living in dwellings that need major repairs

Age and Labour Force Indicators

1. Proportion of the population who are aged 65 and older
2. Dependency ratio (ratio of dependent population: ages 0-14 and 65+ to the working-age population)
3. Proportion of the population not participating in the labor force

Racialized and Newcomer Populations Indicators

1. Proportion of the population who are recent immigrants (arrived in the last 5 years)
2. Proportion of the population who identify as a visible minority

## Notes

### Competing Interest Statement

The authors have declared no competing interest.

### Funding Statement

Institute for Pandemics, University of Toronto

### Author Declarations

University of Toronto IRB

